# Genotype-Phenotype Correlations and Putative Modifier Genes in SYNGAP1 Encephalopathy

**DOI:** 10.1101/2025.10.01.25336001

**Authors:** Selena Aranda, Juliana Ribeiro-Constante, Alba Tristán-Noguero, Nerea Moreno-Ruiz, Concepción Arenas, Fernando Francisco Martínez Calvo, Salvador Ibañez-Mico, José Luis Peña Segura, José Miguel Ramos-Fernández, María del Carmen Moyano Chicano, Rafael Camino León, Víctor Soto-Insuga, Elena González-Alguacil, Carlos Valera Dávila, Alberto Fernández-Jaén, Laura Plans, Ana Camacho, Nuria Visa-Reñé, María del Pilar Martin-Tamayo Blázquez, Fernando Paredes-Carmona, Itxaso Marti-Carrera, Guillem Ginot-Julià, Aránzazu Hernández-Fabián, Meritxell Tomas Davi, Merce Casadesus Sanchez, Laura Cuesta Herraiz, Patricia Fuentes Pita, Teresa Bermejo Gonzalez, Mar O’Callaghan, Federico Felipe Iglesias Santa Polonia, María Rosario Cazorla, María Teresa Ferrando Lucas, Antonio González-Meneses, Júlia Sala-Coromina, Alfons Macaya, Amaia Lasa-Aranzasti, Anna Ma Cueto-González, Francisca Valera Párraga, Jaume Campistol Plana, Mercedes Serrano, Xenia Alonso, Maria Irene Valenzuela Palafoll, Eines Monteagudo, Itziar Alonso-Colmenero, Oscar Sans Capdevila, Ferran Casals, Bru Cormand, Angeles García-Cazorla, Àlex Bayés, Marina Mitjans

## Abstract

Synaptic Ras GTPase-Activating Protein 1 (SynGAP) is a key regulator of synaptic plasticity, neurodevelopment, and neuronal circuit function. It is encoded by the *SYNGAP1* gene, in which *de novo* dominant pathogenic variants are a major cause of SYNGAP1 Encephalopathy, a rare neurodevelopmental disorder characterized by intellectual disability, epilepsy, autistic traits, and other clinical manifestations. While some genetic studies have reported genotype-phenotype correlations in this condition, our understanding of how specific genetic variants contribute to the heterogeneous clinical symptoms remain limited. Here, we analysed a cohort of 44 cases extensively characterised at the phenotypic level to investigate the impact of genetic variants in *SYNGAP1* and in potentially modulatory genes on the clinical features of SYNGAP1 Encephalopathy. Our results indicate that patients with variants in the PH domain of SynGAP exhibited milder phenotypes than other individuals. Moreover, missense variants were associated with a higher prevalence of autistic traits compared to loss-of-function variants. Autistic traits also showed a suggestive positive correlation with the predicted length of the encoded protein. Finally, patients harbouring rare or low-frequency variants in *SYNGAP1*- related genes tended to present with higher global severity. Taken together, these findings suggest that both the location and nature of *SYNGAP1* variants, along with additional genetic modifiers, may contribute to the variability in clinical presentation and severity. Further studies involving larger cohorts and functional validation are needed to refine genotype-phenotype correlations and support the development of personalized management strategies.

## Introduction

The Synaptic Ras GTPase-Activating Protein 1 (SynGAP) is a repressor of small GTPases mainly expressed in forebrain regions (1). It is highly expressed in the postsynaptic density of excitatory glutamatergic neurons, where it regulates the trafficking of the amino-3-hydroxy-5- methyl-4-isoxazolepropionic acid receptor (AMPAR) to the postsynaptic membrane (2), contributing to synaptogenesis, neural circuit function, and synaptic plasticity (1).

SynGAP contains three well-structured N-terminal domains. The Pleckstrin Homology (PH) domain would ensure the membrane recruitment of SynGAP by binding to phospholipids (3), while the C2 and GTPase-activating protein (GAP) domains mediate its GAP catalytic activity (4). Additionally, the protein features a disorganised C-terminal pseudo-domain homologous to Disabled-2-interacting protein (DAB2PC), which includes a Src homology 3 (SH3) binding motif that might mediate the interaction of SynGAP with SH3-domain-containing proteins (5), and a coiled-coil (CC) domain that promotes trimerisation and local concentration of SynGAP (6).

*De novo* variants in *SYNGAP1* are a frequent cause of SYNGAP1 Encephalopathy (7–10), a rare autosomal dominant neurodevelopmental disorder with a prevalence reported as 1:16,000 individuals (11) and characterized by a highly variable clinical presentation. Most individuals with SYNGAP1 Encephalopathy exhibit developmental delay, intellectual disability (ID), and epilepsy (12,13), being autistic traits, severe sleep disturbances, and behavioural problems also common manifestations (1,12–15).

To date, several individual case reports (16–27) and cohort studies (28–36) of patients with SYNGAP1 Encephalopathy have been published. Nevertheless, only a limited number of studies have attempted to establish genotype-phenotype correlations (37,38), and just four have employed statistical analyses to support their conclusions (12,13,39,40). These studies have reported that variants located in exons 1-5, which encode the PH domain, are associated with milder neurodevelopmental delay (39), ID (12), language impairment (40) and epilepsy (13). Also, variants in the SH3 binding motif have been found to be less frequent among patients with epilepsy (39). Despite these insights, a comprehensive understanding of how *SYNGAP1* variants contribute to the variability in clinical features and penetrance remains incomplete. Moreover, several other monogenic neurodevelopmental disorders share clinical features with SYNGAP1 Encephalopathy (41–45), raising the possibility that the observed phenotypic heterogeneity may be influenced by variants in additional genes.

In light of this, we analysed a well-characterized cohort of patients with SYNGAP1 Encephalopathy, comprising both previously reported individuals (36) and newly diagnosed cases, to identify novel genetic variants and establish genotype-phenotype correlations. Furthermore, we investigated whether variants in other genes may modulate clinical severity in this disorder.

## Material and methods

### SYNGAP1 Encephalopathy Cohort

A total of 44 patients diagnosed with SYNGAP1 Encephalopathy were included in this study. Of these, 36 had been previously described (36) while 8 are reported and characterised for the first time in this work. Individuals affected by this condition were recruited through a Spanish network of collaborating child neurologists, geneticists, and psychiatrists, as well as through the *Asociación SynGAP1 España* (www.syngap1.es). Inclusion criteria consisted of (i) a diagnosis of developmental encephalopathy and (ii) the presence of pathogenic or likely pathogenic *SYNGAP1* genetic variants.

Written informed consent was obtained from all parents or legal guardians prior to participation. The study was approved by the local institutional ethics committee (Children’s Hospital Sant Joan de Déu, ID: PIC-232-20) and conducted in accordance with the Declaration of Helsinki.

### Clinical phenotyping

A standardised phenotypic questionnaire was provided to referring physicians to assess seven clinical features: ID, gross motor disabilities, language delay, behavioural abnormalities, autistic traits, epilepsy and sleep disorders (36). For the purpose of this study, each clinical feature was subsequently scored, with higher values indicated greater severity.

ID was determined using Intelligence Quotient (IQ) scores or information regarding functional levels, in accordance with the fifth edition of the *Diagnostic and Statistical Manual of Mental Disorders* (DSM-5) (46). ID severity was categorised into four levels: 1-mild, 2-moderate, 3- severe, and 4-profound.

Gross motor disabilities were evaluated using the Gross Motor Function scale (47), with levels 1 to 3 indicating increasing severity.

Current language ability was assessed during clinical evaluation and categorised into four levels: 1-able to produce complex sentences, 2-able to associate words or simple sentences, 3- able to speak individual words, and 4-absence of speech.

Behavioural abnormalities were assessed across nine questions evaluating aggression, anxiety, flat affect, impulsivity, inattention, hyperactivity, oppositional behaviour, obsessive-compulsive behaviour, and self-injurious behaviour. Each trait was scored as either present or absent, yielding a total score ranging from 0 to 9. For patients with missing data on one to three traits, scores were proportionally scaled to maintain a maximum possible value of 9. Patients with missing data on four or more traits were excluded from the analyses related to behavioural abnormalities.

Autistic traits (poor visual contact, deficits in social communication, restricted and repetitive patterns of behaviour, insistence on sameness, highly restricted and fixated interests, and hyper- or hypo-reactivity to sensory input) were assessed according to DSM-5 criteria for autism spectrum disorder (ASD), although patients did not necessarily fulfil the full diagnostic criteria for ASD. A cumulative score ranging from 0 to 6 represented the total number of traits present. If data were missing for one or two traits, the remaining trait scores were proportionally adjusted to preserve the 6-point maximum score. Patients with missing data for three or more traits were excluded from analyses related to autistic traits.

Epilepsy, when present, was classified as either refractory or non-refractory, depending on whether seizures persisted despite appropriate medication with at least two well-tolerated and properly selected antiseizure drugs at the time of data collection (48). Further subtyping of seizures was not conducted due to the significant phenotypic heterogeneity associated with SYNGAP1-related epilepsy and the limited reliability of clinical data for accurate seizure classification.

Sleep disorders were assessed using the Sleep Disturbance Scale for Children (49).

### Global severity score definition

A global severity score ranging from 0 to 100 was calculated to quantify the overall clinical severity of each patient. This score was derived by summing seven equally weighted sub-scores, each corresponding to one of the clinical features described above. In cases where data for one or more features were missing, the total score was recalculated based on the available features, with weights adjusted to ensure equal contribution and to maintain the 0 to 100 scale.

Sub-scores were computed using feature-specific procedures. For ID, gross motor disabilities, and language delay, values were assigned according to the levels of severity assessed in each patient and proportionally scaled. Absence of the feature resulted in a score of 0. For behavioural abnormalities and autistic traits, sub-scores were calculated by multiplying the number of traits present by the corresponding item weight, defined as the maximum feature score divided by the number of available items for each patient (i.e. 9 for behavioural abnormalities and 6 for autistic traits when complete). For epilepsy, patients with no history of seizures were assigned a sub-score of 0, those with non-refractory epilepsy received half the maximum sub-score, and those with refractory epilepsy received the full score. Finally, sleep disorders were scored in a binary manner: 0 if absent and the full sub-score if any sleep disorder was reported.

### Statistical analysis of sex differences in clinical features

Sex differences in the clinical manifestation of SYNGAP1 Encephalopathy were assessed across all clinical features. The Wilcoxon Ranked-Sum test was used to analyse differences in intellectual disability, gross motor disabilities, language delay, behavioural abnormalities, and autistic traits. Fisher’s exact test was applied to compare the prevalence of epilepsy and sleep disorders between sexes.

### Genotyping and characterisation of SYNGAP1 variants

Identification of *SYNGAP1* variants in the eight newly recruited patients followed previously reported procedures (36). Briefly, genomic DNA was isolated from peripheral blood samples using standard protocols at the participating hospitals. *SYNGAP1* variants were identified using either exome sequencing (Illumina technology) or targeted gene panels for neurodevelopmental disorders. All variants were independently confirmed by Sanger sequencing.

*SYNGAP1* variants were classified as missense, frameshift, nonsense, microdeletions, or intronic and were annotated using established databases and protocols (36). Additionally, variants were grouped based on their predicted impact on SynGAP protein expression: those predicted to result in complete absence of the protein and those that did not. Variants predicted to cause absence of protein included microdeletions spanning the *SYNGAP1* gene and variants meeting the criteria for nonsense-mediated mRNA decay (50).

To estimate the expected length of the SynGAP protein encoded by a mutated gene, we applied the following criteria to the longest human SynGAP isoform reported in InterPro (Q96PV0-1) (51). For variants predicted to result in absence of protein, the protein length was set to zero. For missense variants not predicted to be buried in the protein core, the full protein length (1,343 amino acids) was retained. For loss-of-function variants (nonsense and frameshifts) predicted to skip nonsense-mediated mRNA decay (50), protein length was estimated as follows: for nonsense variants, the length corresponded to the total number of amino acids up (but not including) the premature stop codon introduced by the variant. For frameshift variants, length was calculated as the number of amino acids up to the first downstream stop codon following the frameshift.

### Statistical analyses of SYNGAP1 variant distribution across the gene

To assess whether *SYNGAP1* variants were uniformly distributed across the gene or concentrated within specific functional domains, we compared the observed versus expected frequency of variants within protein domains using the Fisher’s exact test. The same procedure was applied to pathogenic or likely pathogenic *SYNGAP1* variants retrieved from the ClinVar repository, which lists genetic variants identified in clinical samples (52). Expected counts were estimated by multiplying the length of each protein domain by the probability of observing exactly one variant under the assumption of a uniform distribution. This probability was derived from a Poisson model, using the total number of *SYNGAP1* coding variants in our cohort (N=41) and in ClinVar (N=1,376), along with the length of the longest human SynGAP isoform (Q96PV0-1) (51). Variants located within the DAB2PC pseudo-domain were considered outside functional domains due to its unstructured nature.

### Statistical analyses of SYNGAP1 genotype-phenotype correlations

We investigated associations between clinical features and both the type and location of *SYNGAP1* variants using the Kruskal-Wallis test. For analyses based on variant type, we performed the following comparisons: (i) missense versus loss-of-function variants; (ii) missense, frameshift versus nonsense variants; and (iii) variants predicted to result in absence of protein versus those not predicted to do so. For variant location, we compared clinical features between patients with variants located inside versus outside of protein domains or pseudo-domains, as well as between patients with variants located in different specific protein domains or pseudo-domains. Additionally, we evaluated the relationship between the severity of clinical features and the predicted length of the encoded SynGAP protein using Fisher’s exact test or the Spearman’s correlation coefficient, depending on the nature of the data.

### Identification of variants in SYNGAP1-related genes and correlation with clinical features

To explore whether variants in other genes influence clinical features in the 12 patients with available exome sequencing data, we employed two different approaches. First, we investigated whether patients harboured variants in genes encoding proteins with high-confidence interactions with SynGAP, according to STRING (minimum required interaction score = 0.7, maximum number of interactors = 50) (53). We then tested whether patients carrying rare or low-frequency variants (minor allele frequency < 0.05) in these genes had different global severity scores using the Wilcoxon test. Second, we retrieved the total number of variants carried by each patient in genes belonging to eight categories considered relevant to SYNGAP1 Encephalopathy: (i) ID and (ii) epilepsy genes, as defined by the Human Phenotype Ontology Database (54); (iii) brain development and (iv) neuron development genes, based on Gene Ontology Terms ‘Brain Development’ (GO:0007420) and ‘Neuron Development’ (GO:0048666); (v) a SynGAP interactome derived from mouse brain (55); (vi) genes associated with ASD in SFARI (56); (vii) an expert-curated list of synaptic proteins; and (viii) a manually curated list of neurotransmitter receptors and plasma membrane proteins (Table S1). We then tested whether the variant load in each of these functional categories was associated with the presence of ID, autistic traits, epilepsy, and the global severity score.

In all analyses, a P-value (P) less than 0.05 was considered statistically significant. When adjustment for multiple comparisons was required, adjusted P-values (P-adj) were calculated using the Benjamini-Hochberg method.

## Results

### Sociodemographic and clinical features of patients

Male patients represented 54.55% of the studied cohort, with a mean age at diagnosis of 8.64 ± 4.54 years and a mean age at interview of 11.60 ± 5.30 years. Patients were treated with antiseizure drugs (75%), antipsychotics (38.09%), or other medications (34.14%) (Table S2). No statistically significant differences in clinical manifestations were observed between sexes (Table S3).

All 44 patients presented with ID and language delay. The majority also exhibited behavioural abnormalities (97.63%), autistic traits (97.68%), gross motor impairments (83.72%), epilepsy (75%), and sleep disorders (69%) (Fig. 1). Fig. 2A summarizes the phenotypic features and clinical manifestations of the patients, with colour intensity reflecting severity. The global severity score ranged from 22 to 90, with a mean of 60.5 and a coefficient of variation of 17.3% (Fig. 2A, Table S4). Most clinical features were strongly correlated with each other and with the global severity score, with the exception of autistic traits (Fig. 3).

**Figure 1.**
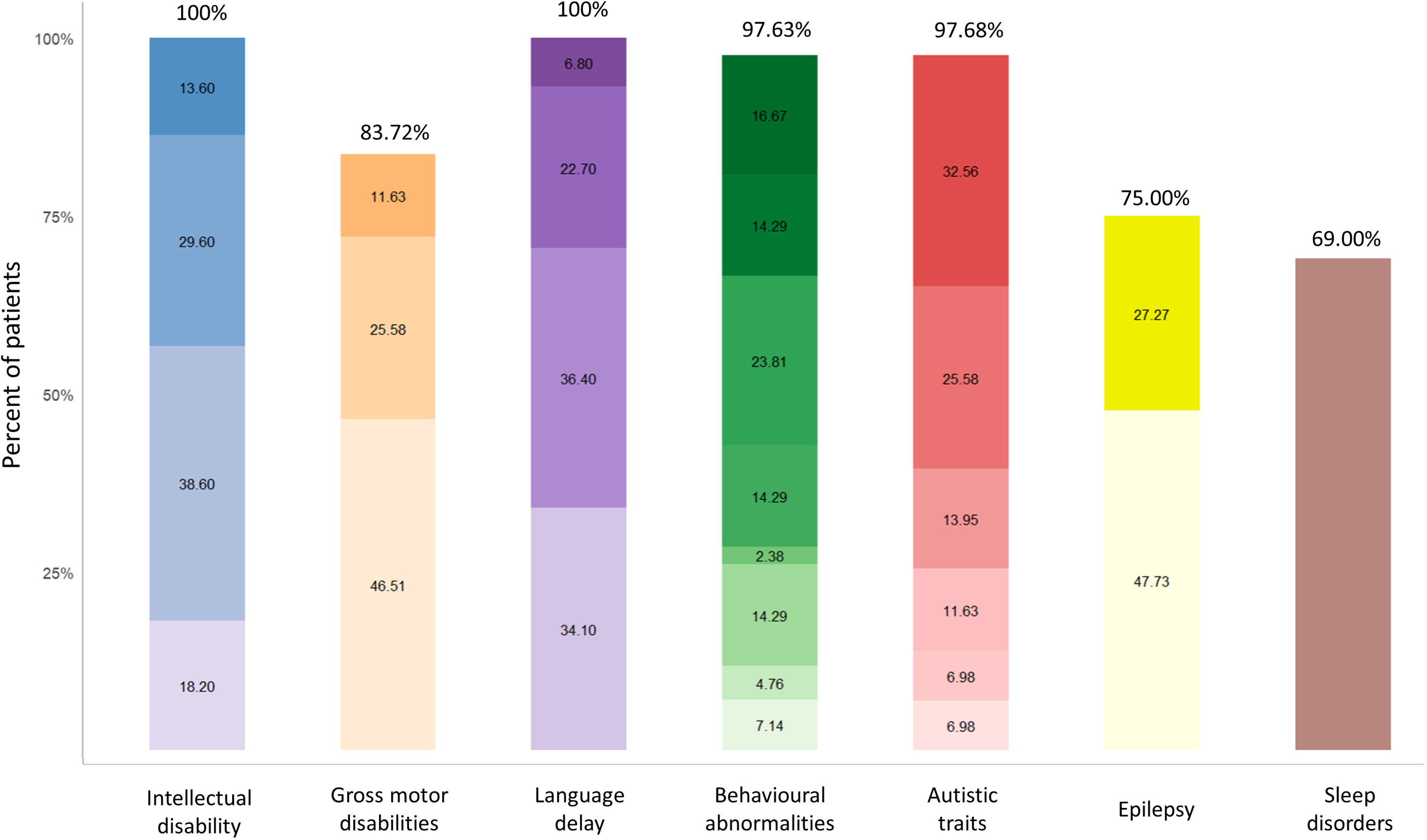
Clinical features of patients. The figure shows the percentage of patients with presenting clinical features. In the first five columns, light to dark colours indicate an increasing level of intellectual disability, gross motor disabilities and language delay; or increasing number of behavioural abnormalities and autistic traits. Light yellow denotes the presence of non- refractory epilepsy; dark yellow denotes the presence of refractory epilepsy. The brown bar indicates the presence of sleep disorders. See Supplementary Table 1 for more details.

**Figure 2.**
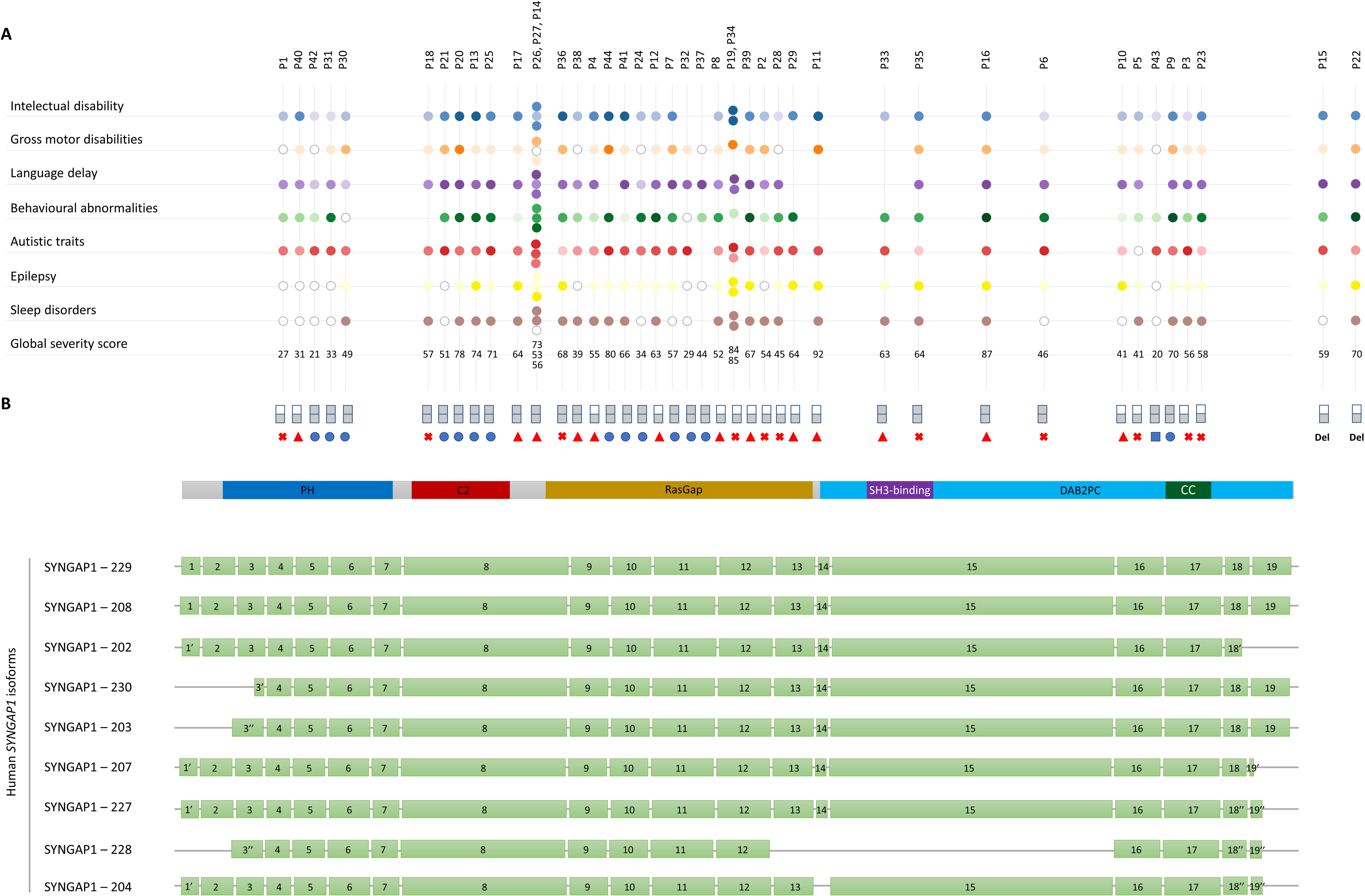
Clinical features of patients depending on the type and location of *SYNGAP1* variants across human isoforms. (A) Phenotypic features and clinical manifestations of the patients. Vertical lines represent individual patients, while horizontal lines represent clinical data. In the first five horizontal lines, colours range from light to dark, indicating increasing levels of intellectual disability, gross motor disabilities, language delay, and an increasing number of behavioural abnormalities and autistic traits. Light yellow denotes the presence of non-refractory epilepsy, while dark yellow represents refractory epilepsy. Brown circles indicate the presence of sleep disorders, and white dots denote the absence of the clinical feature. **(B) Types of mutations and their location across the different human isoforms of the *SYNGAP1* gene.** Genetic variants are represented as blue circles (missense variants), blue squares (intronic variants), red triangles (frameshift variants), and red crosses (nonsense variants). Half-coloured rectangles indicate variants predicted to cause absence of protein, while fully coloured rectangles represent variants that do not affect protein quantity. The grey bar shows the SynGAP1 protein and the domains or pseudo-domains where the variants are located. Green bars represent the coding exons of alternatively spliced SynGAP1 isoforms, according to www.ensembl.org.

**Figure 3.**
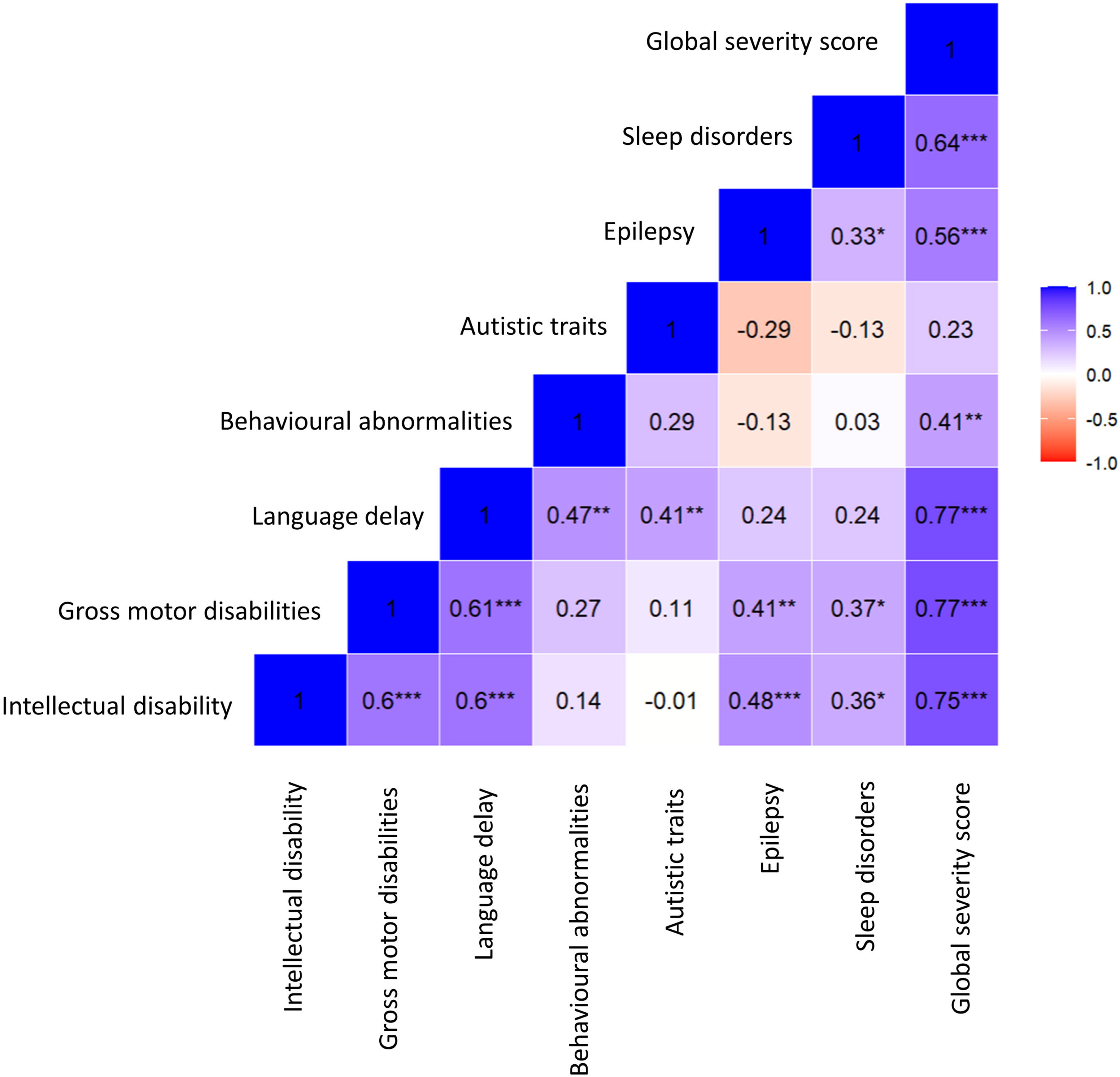
Correlation matrix between clinical features. Each cell displays the Spearman correlation coefficient between two variables. Red colours indicate negative correlation and blue colours indicate positive correlation. The asterisks indicate significance values: ***P-adj ≤ 0.001, **P-adj ≤ 0.01, *P-adj < 0.05.

### Outline of SYNGAP1 genetic variants in the cohort

Among the newly characterised patients, four (P37-40) carried previously reported *SYNGAP1* variants, while other four (P41-44) harboured novel variants, identified for the first time in this study. In the seven patients for whom parental DNA was available, *SYNGAP1* variants were confirmed to have occurred de novo (Table S2).

Across the entire cohort, 27 *SYNGAP1* variants were classified as loss-of-function, including 15 frameshift and 12 nonsense variants. Among these, three cases (P14: c.1171_1172delGG, p.Gly391Glnfs*27; and P26 and P27: c.1167_1168delAG; p.Gly391Glnfs*27) carried a frameshift variant affecting the same codon, and two (P19, P34) shared the same exact nonsense variant (c.1861C>T; p.Arg621*) (Fig. 2B, Table S2). Additionally, 14 variants were classified as missense. Of these, the one from patient P9 is predicted to impact splicing due to its proximity to the splice site (57), and those from patients P7, P13, P20, P21, P24, P25 and P41 were buried in the protein core of SynGAP (0-9% relative solvent accessibility according to PolyView (58)). Finally, the cohort also included 2 microdeletions encompassing the *SYNGAP1* gene (P15 and P22), and 1 intronic variant (P43), classified as likely pathogenic by Varsome (59) and according to the criteria defined by the ACMG (American College of Medical Genetics), as applied in our previous study on this cohort (36) (Fig. 2B, Fig. 4).

**Figure 4.**
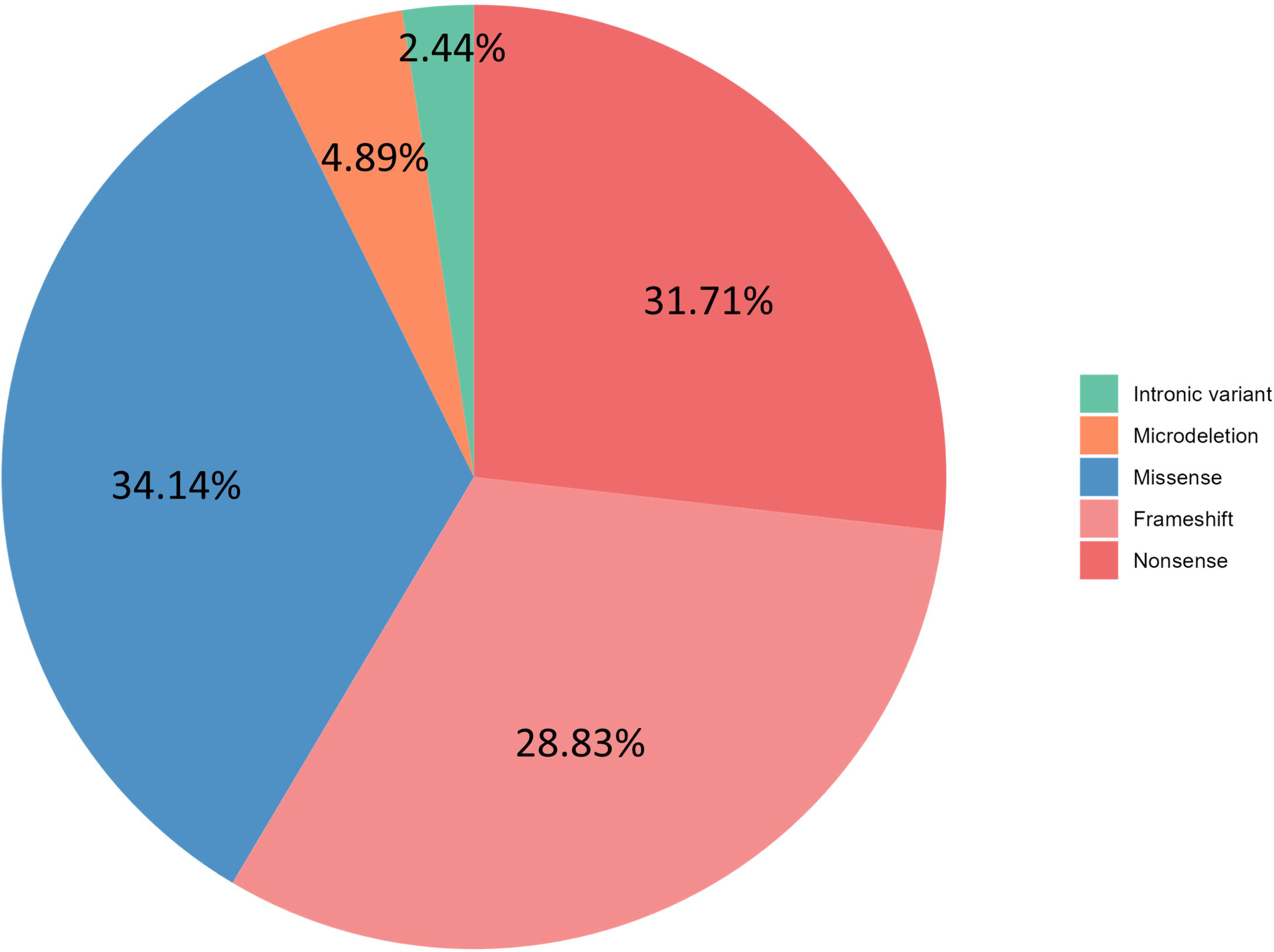
Genetic features of patients. The pie chart shows the frequency of the different types of mutations carried by patients in our sample. Loss-of-function variants are divided into frameshift and nonsense variants.

### Distribution of SYNGAP1 variants

We did not observe an enrichment of *SYNGAP1* variants in the functional domains in our cohort, either when considering the three domains together or when analysing them individually (Table 1). In ClinVar, the number of pathogenic or likely pathogenic variants observed was lower than expected within the PH domain when considering all variants (P = 5.22E-05) and LoF variants (P = 2.20E-04), and higher than expected in the C2 and GAP domains when considering missense variants (P = 0.010 and P = 0.024, respectively) (Table S5). The distribution of pathogenic or likely pathogenic *SYNGAP1* variants in ClinVar is shown in Fig. S1.

**Table 1.** Comparison between the observed and expected mutations in SYNGAP1 functional domains in our cohort.

| Protein domain | All variants |  |  | Missense variants |  |  | Loss of function variants |  |  |
| --- | --- | --- | --- | --- | --- | --- | --- | --- | --- |
|  | N<sub>observed</sub> | N<sub>expected</sub> | P | N<sub>observed</sub> | N<sub>expected</sub> | P | N<sub>observed</sub> | N<sub>expected</sub> | P |
| PH + C2 + GAP | 27 | 21 | 0.179 | 14 | 14 | 1 | 13 | 7 | 0.178 |
| PH | 5 | 7 | 0.532 | 2 | 5 | 0.256 | 3 | 2 | 0.654 |
| C2 | 5 | 4 | 0.724 | 1 | 2 | 0.56 | 4 | 1 | 0.179 |
| GAP | 17 | 10 | 0.100 | 11 | 7 | 0.344 | 6 | 4 | 0.526 |
| Out of domain | 14 | 20 | 0.179 | 13 | 13 | 1 | 1 | 7 | 0.178 |

### SYNGAP1 genotype-phenotype correlations

No statistically significant associations were found between variant types and the severity of clinical features after correction for multiple testing (Table 2). However, patients carrying missense variants showed a trend toward increased autistic traits (P = 0.037, P-adj = 0.296) (Table 2).

**Table 2.** Comparison of the severity of the clinical features depending on the mutation type.

|  | Missense<br>(N=14) | Loss of function<br>(N=27) | P | P-adj |
| --- | --- | --- | --- | --- |
| Intellectual disability (mean ± SD) | 2.79 ± 1.05 | 2.59 ± 0.87 | 0.462 | 0.792 |
| Gross motor disabilities (mean ± SD) | 1.43 ± 0.94 | 1.32 ± 0.86 | 0.710 | 0.802 |
| Language delay (mean ± SD) | 3.00 ± 1.24 | 3.00 ± 0.76 | 0.593 | 0.792 |
| Behavioural abnormalities (mean ± SD) | 5.21 ± 2.52 | 5.22 ± 2.19 | 0.802 | 0.802 |
| Autistic traits (mean ± SD) | **5.21 ± 0.8** | **3.89 ± 1.89** | **0.037** | **0.296** |
| Epilepsy (%) | 64.29 | 82.76 | 0.252 | 0.672 |
| Sleep disorders (%) | 53.85 | 78.57 | 0.146 | 0.584 |
| Global severity score (mean ± SD) | 59.31 ± 19.57 | 62.26 ± 15.52 | 0.594 | 0.792 |

|  | Missense<br>(N=14) | Frameshift<br>(N=15) | Nonsense<br>(N=12) | P | P-adj |
| --- | --- | --- | --- | --- | --- |
| Intellectual disability (mean ± SD) | 2.79 ± 1.05 | 2.67 ± 0.62 | 2.42 ± 1.16 | 0.702 | 0.956 |
| Gross motor disabilities (mean ± SD) | 1.43 ± 0.94 | 1.33 ± 0.9 | 1.27 ± 0.9 | 0.956 | 0.956 |
| Language delay (mean ± SD) | 3.00 ± 1.24 | 3.07 ± 0.8 | 2.75 ± 0.62 | 0.204 | 0.765 |
| Behavioural abnormalities (mean ± SD) | 5.21 ± 2.52 | 5.27 ± 2.37 | 4.9 ± 2.13 | 0.791 | 0.956 |
| Autistic traits (mean ± SD) | 5.21 ± 0.8 | 4.47 ± 1.41 | 3.18 ± 2.32 | 0.100 | 0.765 |
| Epilepsy (%) | 64.29 | 80.00 | 83.00 | 0.659 | 0.956 |
| Sleep disorders (%) | 53.85 | 78.57 | 83.33 | 0.287 | 0.765 |
| Global severity score (mean ± SD) | 59.31 ± 19.57 | 63.38 ± 16.27 | 60.1 ± 16.27 | 0.879 | 0.956 |

|  | Absence of protein<br>(N=18) | Presence of protein<br>(N=26) | P | P-adj |
| --- | --- | --- | --- | --- |
| Intellectual disability (mean ± SD) | 2.61 ± 0.92 | 2.64 ± 0.99 | 0.877 | 0.816 |
| Gross motor disabilities (mean ± SD) | 1.31 ± 0.87 | 1.28 ± 0.89 | 1 | 1 |
| Language delay (mean ± SD) | 2.94 ± 0.77 | 2.91 ± 1.04 | 0.869 | 0.816 |
| Behavioural abnormalities (mean ± SD) | 4.81 ± 3.21 | 5.4 ± 3.50 | 0.739 | 0.985 |
| Autistic traits (mean ± SD) | 3.61 ± 1.75 | 4.52 ± 1.36 | 0.093 | 0.816 |
| Epilepsy (%) | 78.00 | 73.00 | 0.738 | 0.816 |
| Sleep disorders (%) | 76.00 | 64.00 | 0.405 | 1 |
| Global severity score (mean ± SD) | 58.07 ± 17.63 | 55.66 ± 18.29 | 0.993 | 0.816 |

P-adj: P-value adjusted using the Benjamini-Hochberg method.

Similarly, no significant differences in clinical features were observed between patients with variants located within versus outside of a protein domain or pseudo-domain (Table 3). However, patients with variants outside of domains tended to present more frequently with epilepsy than those with variants within domains (P = 0.008, P-adj = 0.064).

**Table 3.** Comparison of the severity of the clinical features depending on the location of the mutation.

|  | Protein domain<br>(N=37) | Out of protein domain<br>(N=4) | P | P-adj |  |  |
| --- | --- | --- | --- | --- | --- | --- |
| Intellectual disability (mean ± SD) | 2.62 ± 0.98 | 2.75 ± 0.50 | 0.555 | 0.740 |  |  |
| Gross motor disabilities (mean ± SD) | 1.39 ± 0.90 | 1.00 ± 0.82 | 0.421 | 0.740 |  |  |
| Language delay (mean ± SD) | 2.95 ± 0.94 | 3.00 ± 0.82 | 0.451 | 0.740 |  |  |
| Behavioural abnormalities (mean ± SD) | 5.20 ± 2.32 | 4.75 ± 2.50 | 0.507 | 0.740 |  |  |
| Autistic traits (mean ± SD) | 4.33 ± 1.79 | 4.75 ± 0.96 | 0.782 | 0.782 |  |  |
| Epilepsy (%) | 72.97 | 100 | 0.008 | 0.064 |  |  |
| Sleep disorders (%) | 71.43 | 75.00 | 0.713 | 0.782 |  |  |
| Global severity score (mean ± SD) | 60.67 ± 17.81 | 64.38 ± 9.28 | 0.253 | 0.740 |  |  |
|  | PH domain<br>(N=5) | Other domains<br>(N=27) | P | P-adj |  |  |
| Intellectual disability (mean ± SD) | 1.80 ± 0.84 | 2.81 ± 0.92 | 0.037 | 0.059 |  |  |
| Gross motor disabilities (mean ± SD) | 0.80 ± 0.84 | 1.46 ± 0.9 | 0.145 | 0.165 |  |  |
| Language delay (mean ± SD) | 1.60 ± 0.55 | 3.15 ± 0.82 | 0.002 | 0.008 |  |  |
| Behavioural abnormalities (mean ± SD) | 3.60 ± 2.88 | 5.46 ± 2.02 | 0.088 | 0.117 |  |  |
| Autistic traits (mean ± SD) | 4.20 ± 0.84 | 4.42 ± 1.75 | 0.407 | 0.407 |  |  |
| Epilepsy (%) | 20.00 | 77.78 | 0.024 | 0.048 |  |  |
| Sleep disorders (%) | 20.00 | 84.00 | 0.011 | 0.029 |  |  |
| Global severity score (mean ± SD) | 35.00 ± 10.13 | 64.89 ± 13.87 | 0.0001 | 0.0008 |  |  |
|  | PH domain<br>(N=5) | C2 domain<br>(N=5) | GAP domain<br>(N=17) | DB2PC pseudodomain<br>(N=10) | P | P-adj |
| Intellectual disability (mean ± SD) | 1.80 ± 0.84 | 3.20 ± 0.84 | 2.82 ± 0.95 | 2.40 ± 0.97 | 0.085 | 0.144 |
| Gross motor disabilities (mean ± SD) | 0.80 ± 0.84 | 1.60 ± 0.89 | 1.31 ± 0.95 | 1.70 ± 0.82 | 0.312 | 0.356 |
| Language delay (mean ± SD) | 1.60 ± 0.55 | 3.40 ± 0.89 | 3.06 ± 0.90 | 3.20 ± 0.63 | 0.013 | 0.035 |
| Behavioural abnormalities (mean ± SD) | 3.60 ± 2.88 | 6.20 ± 0.84 | 5.06 ± 2.24 | 5.78 ± 2.54 | 0.279 | 0.357 |
| Autistic traits (mean ± SD) | 4.20 ± 0.84 | 5.25 ± 0.96 | 4.41 ± 1.77 | 3.9 ± 2.38 | 0.654 | 0.654 |
| Epilepsy (%) | 20.00 | 80.00 | 70.59 | 100 | 0.008 | 0.032 |
| Sleep disorders (%) | 20.00 | 80.00 | 80.00 | 80.00 | 0.090 | 0.144 |
| Global severity score (mean ± SD) | 35.00 ± 10.13 | 69.27 ± 12.41 | 62.73 ± 15.55 | 65.69 ± 16.79 | 0.002 | 0.016 |

P-adj: P-value adjusted using the Benjamini-Hochberg method.

Patients with variants in the PH domain presented significantly lower rates of language delay (P = 0.002, P-adj = 0.008), epilepsy (P = 0.024, P-adj = 0.048), and sleep disturbances (P = 0.011, P-adj = 0.029), as well as lower global severity scores (P = 0.0001, P-adj = 0.0008), compared to the rest of the cohort (Table 3). They also showed a trend toward milder ID (P = 0.037, P-adj = 0.059).

A suggestive positive correlation was observed between the length of the encoded SynGAP1 protein and the presence of autistic traits (r = 0.337, P = 0.027), although this association was not significant after correction for multiple testing (P-adj. = 0.216) (Table 4).

**Table 4.** Correlation of the severity of the clinical features with the predicted length of the encoded SynGAP1.

|  | Correlation coefficient | P | P-adj |
| --- | --- | --- | --- |
| Intellectual disability (mean ± SD) | 0.020 | 0.897 | 0.897 |
| Gross motor disabilities (mean ± SD) | 0.044 | 0.776 | 0.897 |
| Language delay (mean ± SD) | 0.056 | 0.719 | 0.897 |
| Behavioural abnormalities (mean ± SD) | 0.103 | 0.514 | 0.897 |
| **Autistic traits (mean ± SD)** | **0.337** | **0.027** | **0.216** |
| Epilepsy (%) | - | 0.282 | 0.752 |
| Sleep disorders (%) | - | 0.121 | 0.484 |
| Global severity score (mean ± SD) | -0.036 | 0.817 | 0.897 |

P-adj: P-value adjusted using the Benjamini-Hochberg method.

Among patients with recurrent variants, the three patients carrying a frameshift mutation affecting the same *SYNGAP1* codon exhibited variable clinical presentations, while the two patients harbouring the same nonsense variant showed a more similar phenotype (Fig. 2A, Table S2).

### Genotype-phenotype correlations for SYNGAP1-related genes

A total of 28 high-confidence SynGAP1 interactors were retrieved from STRING (Fig. S2), and 11 variants in the genes encoding these interacting proteins were identified in the 12 patients with available exome sequencing data in our sample. Most of these variants were rare or of low frequency and occurred at highly conserved sites, with Combined Annotation Dependent Depletion (CADD) scores above 20. Specifically, two patients carried missense variants in *SHANK3* (P25) and *NLGN2* (P16) with a population frequency of 3%; two patients (P8, P31) carried a missense variant in *SHANK3* with a frequency of 1%; three patients had missense variants with a frequency below 0.1% in *DLGAP1* (P9), *NLGN4X* (P16) and *SOS1* (P25); three patients shared a previously undescribed variant in *SHANK1* (P7, P12, P8); and one patient had a frameshift variant in *SHANK1* (P12).Patients harbouring rare or low-frequency variants in SynGAP1-interacting genes showed higher global severity scores (mean ± SD = 65.76 ± 16.92) than those without such variants (mean ± SD = 53.17 ± 15.22), although the difference did not reach statistical significance (P = 0.202) (Table S6). Interestingly, the two patients with the highest global severity scores each harboured two rare or low-frequency variants (Table S6).

Finally, we observed substantial variability in variant burden of the eight compiled gene sets relevant to SYNGAP1 Encephalopathy across patients. However, no evidence of a relationship between variant load in these functional categories of genes and corresponding clinical features was observed (Fig. S3-S9).

## Discussion

In this study, we identified novel genetic variants in *SYNGAP1* and established genotype-phenotype correlations in a phenotypically well-characterised cohort of individuals with SYNGAP1 Encephalopathy. Additionally, we explored the potential contribution of variants in other genes to the observed clinical heterogeneity, providing a broader understanding of the complexity of this disorder.

All patients in our cohort exhibited ID and language delay, while gross motor disabilities and behavioural abnormalities were highly frequent, in line with previous reports (1,12,13,32). The prevalence of epilepsy and sleep disorders was slightly lower, also resembling previously described cohorts (12,13,15). However, the higher frequency of autistic traits found in our cohort (97.68%) contrasts with the prevalence of approximately 50% reported in the literature (12,13), suggesting that these may have been underestimated in SYNGAP1 Encephalopathy. In this regard, in patients with intellectual disability—especially when it is severe—autistic traits can be “masked” by the overall cognitive impairments and be less reported in clinical descriptions. We also report, for the first time, a global severity score to quantify functional impairment, and found that it significantly correlated with all assessed clinical features except for autistic traits. This suggests that the mechanisms underlying autistic traits may be at least partially independent from those influencing other clinical outcomes, although the difficulty to assess the severity of autistic traits in patients with profound intellectual disabilities should not be discarded.

We observed that ID, gross motor disabilities, and language delay were highly correlated to each other. Given the known dependence of cognitive and language development on motor skill progression (60), which is even more pronounced in children with intellectual and developmental disabilities (63), our data emphasises the importance of early motor interventions to eventually enhance cognitive and language development. This is especially relevant in SYNGAP1 Encephalopathy, where motor and language difficulties are often more severe than in other forms of ID (14). Moreover, ID, gross motor disabilities and language delay showed the strongest correlation with the global severity score, being thus their main drivers, and further suggesting that improvements in motor function may lead to broader benefits in overall functioning. One aspect to be further studied in the future is whether different types of motor dysfunction (spastic, dyskinetic, ataxic, mixed) may exhibit distinct trajectories relative to other neurodevelopmental features (cognition, language and behaviour).

The majority of patients in our sample carried loss-of-function variants, followed by missense changes, microdeletions, and intronic variants, mirroring the distribution of *SYNGAP1* variants reported in previous cohorts (12,26,27), and thus corroborating that the ratio of missense to loss-of-function variants in SYNGAP1 Encephalopathy is close to 2:1. In contrast, the annotated variants from the general population in ClinVar show a predominance of missense and intronic variants (52), indicating that missense variants are less likely to cause SYNGAP1 Encephalopathy.

Among the recurrent variants identified in our cohort, the nonsense variant c.1861C>T (p.Arg621*) has been previously reported (61) and occurs at a classical mutational hotspot (CpG > TpG), which may explain its recurrence. In contrast, one of the frameshift variants affecting the same codon, c.1167_1168delAG (p.Gly391Glnfs*27), although not located in a known mutational hotspot, has also been reported (62), further supporting its pathogenic relevance.

The eight newly described patients carried variants mostly located in the PH domain (one missense and one frameshift variant) and the GAP domain (three missense and two frameshift variants). Additionally, one of them carried a variant in intron 16, within a region included in the *SYNGAP1* transcript when a cryptic acceptor splice site is used. This inclusion disrupts the reading frame and results in a prematurely truncated protein product (63).

We found that the number of observed *SYNGAP1* variants in our cohort was higher than expected in the C2 and particularly the GAP domain; however, this difference was not statistically significant. These findings contrast with previous studies suggesting that SYNGAP1 Encephalopathy is primarily driven by dysfunction in the C2 and GAP domains (12,13,64,65). Intriguingly, in ClinVar, we observed fewer pathogenic or likely pathogenic LoF variants than expected within the PH domain and higher pathogenic or likely pathogenic missense variants in the C2 and GAP domains. Taken together, these findings might suggest that while the proper functioning of the C2 and GAP domains is key to prevent SYNGAP1 Encephalopathy, variants in the PH domain may also contribute to the disease.

Our genotype-phenotype analysis revealed that patients with variants in the PH domain exhibited milder phenotypes, including less severe ID, language delay, epilepsy, sleep disorders, and a lower global severity score. These results replicate previous findings indicating that variants in the PH domain are associated with milder ID (12), less severe language delay (66) and less epilepsy (67,68) than patients with variants in other domains. For the first time, we also suggest that such variants may be associated with fewer sleep disturbances and lower global clinical severity. However, the association between PH domain variants and lower presence of epilepsy requires further investigation, as previous reports have also found associations with increased epilepsy risk (12,66,69).

Interestingly, in mice, three SynGAP N-terminal variants (A, B and C) have been described, each influencing synaptic function in a different way (70). Isoforms with a ‘C’ N-terminus, which lacks exons 1-5 encompassing half of the PH domain, increase the amplitude and frequency of excitatory postsynaptic currents with respect to isoforms presenting A and B N-terminals, which retain an intact PH domain (70). Although no human *SYNGAP1* isoforms described to date lack the PH domain (www.ensembl.org), their existence is possible based on rodent findings (70). It is thus plausible that some isoforms starting after the variants in the PH domain are normally expressed in humans, preserving their ability to regulate synapse activity and resulting in milder clinical phenotypes.

Intriguingly, we observed that patients with missense variants exhibited more autistic traits than those with loss-of-function variants, and that autistic traits showed a suggestive positive correlation with the predicted length of the encoded protein. These results suggest that the presence of an aberrant protein may contribute to autistic phenotypes. Although this mechanism has not yet been described in SYNGAP1 Encephalopathy, similar processes have been reported in other neurological conditions involving protein aggregation (71), cytosolic accumulation (72), or disruption of cellular pathways (73). The role of aberrant SynGAP protein forms remains to be demonstrated; however, SynGAP indirectly regulates signalling pathways involved in protein synthesis, and it has been proposed that *SYNGAP1* variants may disrupt synaptic proteostasis, contributing to autistic phenotypes (74).

We did not observe differences in clinical features between patients with and without predicted absence of protein, challenging the notion that SYNGAP1 Encephalopathy may be solely driven by haploinsufficiency, as previously suggested (35).

The three patients carrying a frameshift variant in the same codon exhibit different clinical manifestations, suggesting that other factors, including variants in genes other than *SYNGAP1*, may modulate their symptomatology. In contrast, the two patients with the same nonsense variant display more uniform symptoms and severity, indicating a possible absence of modulatory factors. However, exome sequencing data that could help support this hypothesis are unavailable for these patients.

Finally, among the patients for whom exome sequencing data were available, those harbouring rare or low-frequency functional genetic variants in the *SYNGAP1*-related genes *SHANK3*, *SOS1*, *NLGN2*, *NLGN4X* and *DLGAP1* had a higher global severity score than the other patients. These genes encode postsynaptic proteins implicated in neurodevelopmental disorders (75–79). Notably, knockdown of *SHANK3* in zebrafish produces disruptions in the nervous system that are similar to those observed with *SYNGAP1* knockdown (80), suggesting functional convergence. In this sense, we noticed that the only patient harbouring a missense variant in the DAB2PC pseudo-domain (P9) also had a rare variant in *DLGAP1*, a gene linked to ASD, ID and epilepsy (79). This co-occurrence may explain the unexpectedly high global severity score found in patient P9, holding a missense variant located in a typically less impactful domain.

In summary, our findings suggest that the clinical features of SYNGAP1 Encephalopathy are primarily driven by alterations in the function of the C2 and GAP domains, while autistic traits may be influenced by the presence of an aberrant form of the SynGAP protein. We also provide evidence that variants in related genes may modulate disease severity. We propose that early neuropsychological interventions targeting motor development, combined with targeted pharmacological approaches aimed at specific *SYNGAP1* isoforms, could help mitigate overall disease severity.

## Data Availability

All data produced in the present study are available upon reasonable request to the authors.

## Author contributions

SA: Conceptualization, Investigation, Validation, Visualization, Writing–original draft. JR-C: Conceptualization, Data curation, Validation, Writing–review and editing. AT-N: Conceptualization, Investigation, Writing–review and editing. NM-R: Investigation, Writing– review and editing. CA: Investigation, Writing–review and editing. FM, SI-M, JP, JR-F, MMo, RC, VS, EG, CV, AF-J, LP, AC, NV-R, MM-T, FP-C, IM-C, AH-F, MT, MC, LC, PF, TB, MO’, FI, MR, MF, AG-M, JS-C, EM, AL-A, AC-G, FV, JC, MS, XA, MV, EM, IA-C, OS-C: Resources, Writing–review and editing. GG-J: Investigation, Writing-review and editing. FC: Conceptualization, Investigation, Writing-review and editing. BC: Conceptualization, Investigation, Writing-review and editing. AG-C: Conceptualization, Data curation, Investigation, Writing-review and editing. AB: Conceptualization, Investigation, Writing-review and editing. MMi: Conceptualization, Investigation, Writing–original draft, Writing-review and editing.

## Funding

The authors declare that financial support was received for the research, authorship, and/or publication of this article. AG-C is supported by FI21/0073 and FI24/00469 “Instituto de Salud Carlos III (ISCIII)” and “Fondo Europeo de desarrollo regional (FEDER)”. AT-N was supported by a “Margarita Salas” contract from Next-Generation Europe and postdoc mobility grants “EMBO short exchange research” grant and “José Castillejo” grant. AB financial support was provided by: PID2024-160538OB-I00, PID 2021-124411OB-I00 and RTI 2018-097037-B- I00 (MINECO/MCI/AEI/FEDER, EU), Award AC17/00005 by ISCIII through AES2017 and within the NEURON framework, Ramón y Cajal Fellowship (RYC-2011-08391p), IEDI-2017- 00822 and AGAUR (2017 SGR 1776 and 2021 SGR 01005). AB and AT-N thank the CERCA Programme/Generalitat de Catalunya for institutional support. NM-R was supported by grant 2021 FI_B_00296 from Agència de Gestió d’Ajuts Universitaris i de Recerca, Generalitat de Catalunya. AC-G is member of the European Reference Network on Rare Congenital Malformations and Rare Intellectual Disability ERN-ITHACA, funded by the European Union, under the grant agreement N°101156387. The study was supported by the generous funding provided by the association of Spanish SYNGAP1-DEE families, SYNGAP1 España. AB and BC wish to thank support from the Spanish Red de Investigación RED2024-154082-T.

Funding supporting this study was provided by the Spanish ‘Ministerio de Ciencia, Innovación y Universidades’, funded by MICIU/AEI/10.13039/501100011033/ and FEDER-EU (PID2021- 1277760B-I00 and PID2024-158634OB-I00, to BC; PID2022-139740OA-I00 to MMi; PID2021-125106OB-C32 to FC), ‘Generalitat de Catalunya/AGAUR’ (2021-SGR-01093, to BC and MMi), ICREA Academia 2021 (to BC), and ‘Fundació La Marató de TV3′ (202218-31, to BC). This article is part of the grant RYC2021-033573-I funded by MCIN/AEI/10.13039/501100011033 and by the European Union “NextGenerationEU”/PRTR”. SA is supported by a Juan de la Cierva fellowship from the Spanish Ministry of Science and Innovation (JDC2024-055161-I).

## Conflict of interest

AG-C has received honoraria for research support and lectures from PTC Therapeutics, she has received honoraria for lectures from Biomarin, Immedica and Recordati Rare Diseases Foundation, and is a co-founder of the Hospital Sant Joan de Déu start-up “Neuroprotect Life Sciences”.

The remaining authors declare that the research was conducted in the absence of any commercial or financial relationships that could be construed as a potential conflict of interest.

## Acknowledgements

This work has been made possible through the collaboration of neurologists from across Spain and the invaluable support of the Asociación SynGAP1 España, to whom we extend our heartfelt gratitude. We also sincerely thank all the parents and caregivers, on behalf of the patients, for their participation in this study.

## Notes

### Author Declarations

Ethics committe of Children Hospital Sant Joan de Deu, ID: PIC-232-20, gave ethical approval for this work.

